# Acute intermittent hypoxia-induced increases maximal motor unit discharge rates in people with chronic incomplete spinal cord injury

**DOI:** 10.1101/2023.05.22.23290235

**Authors:** Gregory E P Pearcey, Babak Afsharipour, Aleš Holobar, Milap S Sandhu, W Zev Rymer

## Abstract

Acute intermittent hypoxia (AIH) is an emerging technique for enhancing neuroplasticity and function in respiratory and limb musculature. Thus far, AIH-induced improvements in strength have been reported for upper and lower limb muscles after chronic incomplete cervical spinal cord injury (iSCI) but the underlying mechanisms have been elusive. We used high-density surface electromyography (HDsEMG) to determine if motor unit discharge behaviour is altered after 15 × 60 s exposures to 9% inspired oxygen interspersed with 21% inspired oxygen (AIH), compared to breathing only 21% air (SHAM). We recorded HDsEMG from the biceps and triceps brachii of seven individuals with iSCI during maximal elbow flexion and extension contractions, and motor unit spike trains were identified using convolutive blind source separation. After AIH, elbow flexion and extension torque increased by 54% and 59% from baseline (p = 0.003), respectively, whereas there was no change after SHAM. Across muscles, motor unit discharge rates increased by ∼4 pulses per second (p = 0.002) during maximal efforts, from pre to post AIH. These results suggest that excitability and/or activation of spinal motoneurons are augmented after AIH, providing a mechanism to explain AIH-induced increases in voluntary strength. Pending validation, AIH may be helpful in conjunction with other therapies to enhance rehabilitation outcomes due to these enhancements in motor unit function and strength.

## INTRODUCTION

Acute intermittent hypoxia (AIH; i.e., repeated but brief periods of breathing low oxygen gas mixtures) has facilitatory effects on respiratory and non-respiratory motor function (Sandhu & Rymer, 2021; Vose *et al*., 2022). In reduced preparations, it has been shown that AIH triggers the release of serotonin (5-HT), which is a potent neuromodulator of spinal motoneurons [for reviews, see: (Perrier *et al*., 2013; Perrier & Cotel, 2015; Kavanagh & Taylor, 2022)], and is associated with increases in the synthesis of brain-derived neurotrophic factor (BDNF), one of the most important regulators of neuroplasticity (Baker-Herman *et al*., 2004; Lovett-Barr *et al*., 2012; Sandhu & Rymer, 2021). Our lab has shown that a single sequence of AIH (i.e., 30 minutes) can significantly increase function in the somatic motor system of individuals living with chronic incomplete spinal cord injury, which is evidenced by increased strength of muscles acting at the ankle joint (Trumbower *et al*., 2012; Lynch *et al*., 2017; Sandhu *et al*., 2019), improved wrist/hand function (Sandhu *et al*., 2021), and increased elbow flexor and extensor strength (Afsharipour *et al*., 2022). Despite the known mechanisms responsible for AIH-induced neuroplasticity in the neural control of respiratory muscles observed in several studies by Mitchell and colleagues (Baker & Mitchell, 2000; Baker-Herman & Mitchell, 2002; Baker-Herman *et al*., 2004; Golder & Mitchell, 2005; Vinit *et al*., 2009; Lovett-Barr *et al*., 2012; Satriotomo *et al*., 2016; Dale *et al*., 2017), little is known about the mechanisms responsible for AIH-induced neuroplasticity in human muscle function. The aim of the current investigation was to probe mechanisms underlying AIH-induced increases in strength.

AIH-induced changes in the amplitude of bipolar surface electromyograms (sEMG) recorded from plantar flexor muscles of the ankle during *maximal* plantar flexion contractions have been reported in previous studies (Trumbower *et al*., 2012; Sandhu *et al*., 2019). Additionally, the changes in spatial distribution and increased intensity of high-density surface EMG (HDsEMG) of the biceps and triceps brachii during *maximal* isometric elbow flexion and extension contractions suggests that activation and/or excitability of alpha motoneurons may be enhanced by AIH. Indeed, corticospinal synaptic plasticity is enhanced after AIH (Christiansen *et al*., 2018), but other central nervous system sites may also contribute to the observed functional adaptations.

The quantal elements for force generation are the motor units (Heckman & Enoka, 2012), which are comprised of a spinal motoneuron and all its innervated muscle fibers. Serotonin has potent effects on spinal motoneurons in two ways: 1) directly by affecting afterhyperpolarization (AHP) amplitude and duration; and 2) indirectly through the facilitation of voltage-dependent persistent inward currents (PICs). Both mechanisms could enhance motoneuronal discharge rates, which may result in enhanced force output from the muscle since modulation of muscle force occurs via two primary mechanisms: 1) recruitment of previously inactive motor units, and/or 2) increased discharge rate of active motor units (Milner-Brown *et al*., 1973; Heckman & Enoka, 2012).

Our previous HDsEMG investigation of AIH (Afsharipour *et al*., 2022) prompted us to question whether AIH-induced increases in EMG activity were due to recruitment of new motor units or to increased discharge rates of previously active motor units. The lack of clear shape changes in the EMG maps led us to believe that increased EMG amplitudes of the active region probably favors an increase in discharge rates of already active motor units as the leading mechanism. Using advanced decomposition techniques (i.e., blind source separation (Holobar *et al*., 2009*a*; Negro *et al*., 2016)) we performed secondary data analysis of a previous HDsEMG dataset (Afsharipour *et al*., 2022) to track individual motor units from pre-to post-AIH, and to shed light on the underlying mechanisms contributing to the observed increases in strength after AIH.

The purpose of this investigation was to determine if motor unit discharge rates change after AIH. We hypothesized that AIH would enhance motor unit discharge rates in the biceps brachii and triceps brachii during *maximal* voluntary isometric elbow flexion and extension, leading to increased volitional strength in these muscles.

## METHODS

### Participants

Seven individuals (48.1 ± 11.79 years; 5 males, 2 females) with incomplete spinal cord injuries at the cervical level (i.e., C4-C7) volunteered for this study (see table 1 for characteristics of study participants). Participants provided written informed consent to the experimental procedures, which were approved by the local ethics committee at Northwestern University and performed in accordance with the Declaration of Helsinki (IRB protocol STU00201602). The effects of AIH on peak force and surface EMG amplitudes from this dataset have been published elsewhere (Afsharipour *et al*., 2022).

**Table 1.** Participant characteristics.

| **Subject** | **Sex** | **AIS** | **Level** | **Time Since Injury (years)** |
| --- | --- | --- | --- | --- |
| s01 | M | D | C5 | 3 |
| s02 | M | C | C4-C7 | 11 |
| s03 | M | D | C4-C7 | 4 |
| s04 | F | C | C4-5 | 10 |
| s05 | M | D | C4-C5 | 3 |
| s06 | M | C | C5-C6 | 16 |
| s07 | F | D | C5-C7 | 2 |

### Experimental Protocol

Participants underwent both AIH and SHAM AIH sessions in a randomized order, with a minimum washout period of 7 days between the two sessions to avoid carryover effects. The AIH intervention involved 15 exposures to 9% O_2_ for 1 minute each, alternating with 1-minute exposures to 21% O_2_ (normoxia). In contrast SHAM AIH involved alternating exposures to normoxia alone. The participants were blinded to the intervention (i.e., AIH or SHAM AIH) at the time of experiments.

During each session, participants were seated comfortably in a Biodex chair, with the elbow in 120° flexion, shoulder in 30° flexion and 35° abduction, and the forearm pronated to 45°. The forearm was casted in a custom-built fixture centered at the wrist to ensure force isolation. The fixture was equipped with a six degree-of-freedom load cell (Delta from ATI, NC, USA) to record forces generated at the wrist as a result of elbow flexion/extension. Force signals were analog to digitally converted (Power1401, Cambridge Electronic Design Limited, UK) and acquired at a rate of 2000 Hz (Spike2, Cambridge Electronic Design Limited, UK). During elbow flexion and extension, we recorded HDsEMG data using a grid of surface electrodes applied to biceps brachii (BB; long and short heads), and triceps brachii (TB) muscles, respectively. Force and HDsEMG data were synchronized by using a digital trigger.

Participants performed *maximal* voluntary contractions while recording high density surface electromyograms (HDsEMG) signals before (PRE) and 60 minutes after (POST) each intervention. Three *maximal* isometric contractions (MVCs) of elbow flexion and extension were performed for ∼5 s at each timepoint separated by 2 minutes of rest. Investigators monitored real-time visual feedback about participant force trajectories (i.e. resultant force direction and magnitude) and provided verbal encouragement to participants to ensure the intended actions were performed.

### Acute intermittent hypoxia (AIH) intervention

After force and HDsEMG recordings at baseline (i.e., PRE) in both sessions, we secured a latex-free full non-rebreather mask onto the participant using a custom neoprene head strap (Hypoxico INFO). The AIH intervention was administered using a hypoxia generator (Model HYP-123, Hypoxico Inc., New York, NY, USA), which provided 60 s of 9% O_2_ (Fraction of inspired air [FIO_2_]: 0.09) interleaved with 60 s of 21% O_2_ (FIO_2_: 0.21). We repeated the delivery of hypoxia and normoxic air mixtures 15 times per session, for a total of 30 min. Continuous pulse oximetry (Smith Autocorr Plus, Smiths Medical, Dublin, OH, USA) was used to ensure oxygen saturation (SpO_2_) fell to a nadir between 80 and 87% during hypoxic exposures. The SHAM intervention consisted of alternating exposures of normoxic air (i.e. 30 minutes of 21% O_2_).

### Safety monitoring

Cardiorespiratory function (GE Dash 4000 Monitor, Chicago, IL, USA) was monitored throughout the protocols to ensure that heart rate (50–160 beats per min), systolic blood pressure (85–160 mm Hg), and oxyhemoglobin saturation (>75%) remained within safe limits. We also monitored and asked for self-report of any headaches, chest pain, lightheadedness, dizziness, respiratory distress, muscle spasms, and autonomic dysreflexia, before, during, and after treatment.

### High Density Surface Electromyography (HDsEMG)

Before electrode placement, the skin regions for the grid electrodes and reference electrodes were lightly abraded and cleaned with alcohol pads and with abrasive paste (NuPrep, Weaver and Company, Colorado USA) to increase signal to noise ratio and a reference electrode was placed over the lateral epicondyle. After securing the arm, we placed 64-channel HDsEMG grid electrodes arranged in 8 rows and 8 columns (8×8 electrode grid) equally spaced with an 8.5 mm inter electrode distance, over the BB and TB muscles. The grid electrode was attached to the skin overlying the muscle with double sided adhesive tape, and conductive gel was used to minimize electrode impedance. One grid (8×8) was used for BB recording and one grid was dedicated to collect signals from TB. To ensure consistent placement on the BB across participants, we drew a line from the acromion and the distal biceps tendon insertion, which divided the BB into medial-lateral segments and column 4 was placed along this line. We then found the proximal-to-distal midpoint of the muscle and placed the grid such that the central row of the grid (row 4) divided the grid into two equal proximal and distal portions. For TB, we drew a line between the acromion and the medial epicondyle, which again divided the grid into medial and lateral, and placed column 4 over this line. We then found the proximal-to-distal midpoint and placed the central row of the grid (row 4) at this point. The first half of the rows (counting from top) in the grid were considered proximal (above the mid-point), while the other half covered the distal portion (below the mid-point) for both the BB and TB. Electrode grids remained in the same position the before and after the AIH intervention.

A 128-channel Refa (TMSi; Oldenzaal, NL) recording system with a fixed gain (26.5 times) and sampling frequency of 2000Hz was used to collect HDsEMG signals. The electrode grids remained secured and in the same position throughout the PRE and 60min POST intervention timepoints. All signals were recorded in monopolar configuration, where the instantaneous amplitude of each electrode was measured relative to the lateral humeral epicondyle.

### Data processing

All recorded data (Force and HDsEMG) were converted offline to MATLAB (MATLAB R2021a; MathWorks Inc., MA) for further processing. The resultant force was calculated using the axial components from the principal directions sensed by the load cell and the resultant elbow contraction forces were calculated through the vector addition rule. The resultant force signal was then filtered (fourth-order, zero-lag butterworth filter: 0.1–10 Hz).

All HDsEMG decomposition, MU filter transfers, and manual inspection was performed by an experienced investigator (AH) who was blind to the intervention received by participants in each condition. Monopolar HDsEMG signals were initially bandpass filtered (fourth-order, zero-lag butterworth filter: 20–500 Hz) and all channels exhibiting poor signal-to-noise ratio, movement artifacts, poor contact between skin-electrode, or any peculiarities were removed (typically < 5% of all channels). HDsEMG signals were first concatenated across contractions within the same timepoint (i.e., 2 x ∼5s contractions PRE separately from 2 x ∼5s contractions 60min POST; total of ∼10s of HDsEMG signals from each of the timepoints). Once the HDsEMG signals were concatenated, the two contractions PRE and two contractions 60min POST were then independently decomposed into MU pulse trains using the Convolution Kernel Compensation (CKC) algorithm (Holobar & Zazula, 2008; Holobar *et al*., 2009*b*; Holobar & Farina, 2014), and only MUs with pulse-to-noise ratios (PNR) ⩾ 28 dB were kept for further analysis (Holobar *et al*., 2014). Signals obtained from the two contractions at PRE and the two contractions 60min POST were then concatenated and common MUs from each timepoint were identified using the previously proposed MU filter transfer method (Frančič & Holobar, 2021).

During this procedure, MU filters of identified MUs were independently estimated from the PRE and 60min POST timepoints and then transferred in both directions, from PRE to 60min POST and from 60min POST to PRE timepoints, securing the highest possible number of tracked MUs. In this step of MU filter transfer, MU filters from one timepoint were applied to HDsEMG signals from the other timepoint, identifying the MU spike train of the corresponding MU. MU spike trains were then segmented into MU firing patterns using the spike segmentation routine built in the CKC algorithm (Holobar & Zazula, 2008; Holobar *et al*., 2009*b*, 2014). Finally, firing patterns of identified MUs were mutually compared and MU spike trains sharing at least 30% of firings (firing identification tolerance set to 0.5 ms) were considered duplicates of the same MU. Among them, only one spike train with the highest PNR value was retained for further analysis whereas all the other spike trains were discarded.

In addition, only MUs that were identified in at least one PRE and one 60min POST contraction were kept for further analysis. The results of automatic spike segmentation were finally inspected manually and then edited using common procedures that have been described extensively (Del Vecchio *et al*., 2020; Hug *et al*., 2021). In particular, by using the DEMUSE tool software (version 6.0, University of Maribor, Slovenia), MU firings corresponding to interspike intervals of length < 25 ms or > 250 ms (instantaneous frequency > 40Hz or <4 Hz respectively) were visually inspected and either deleted or validated by the expert. Afterwards, instantaneous discharge rates of each MU were determined by computing the inverse of the interspike interval of each MU spike train and smoothed using support vector regression with a custom-written MATLAB script (Beauchamp *et al*., 2022). The mean discharge rate around the maximal force from each MU during each contraction (± 500ms) was obtained from the smoothed spike trains using custom written MATLAB scripts. Any MU with multiple interspike intervals of >250ms during the intended hold phase was discarded from subsequent analysis because this long interval indicates that they were not engaged in repetitive discharge.

### Statistical analysis

We used a randomized, double-blinded crossover design to test our hypotheses that changes in MU discharge rate would predict AIH-induced increase in volitional elbow strength. All statistical analyses were performed using R Statistical Software (v4.1.0; R Core Team 2021). To determine whether force output and mean discharge rates were predicted by the fixed effects of condition, time, muscle, and their interactions we used linear mixed effects models with a random intercept for each trial, participant, and each motor unit in the case of mean discharge rates only (lmer R package; v1.1.27.1; (Bates *et al*., 2015)). To determine significance, we applied Satterthwaite’s method for degrees of freedom (lmerTest R package; v3.1.3; (Kuznetsova *et al*., 2017)). Estimated marginal means were computed (emmeans R package, v1.8.0, (Lenth *et al*., 2022)) and effect sizes were calculated to determine the standardized magnitude of the effect of AIH from the estimated marginal means of PRE and 60min POST data from the model.

## RESULTS

### Motor unit decomposition during maximal contractions

In the biceps brachii, we decomposed and tracked a total of 95 MUs from pre- to post-AIH (13.6 ± 4.28 per participant; range of 9-19), while we also decomposed and tracked 70 MUs from pre- to post-SHAM (10 ± 7.12 per participant; range of 2-23). In the triceps brachii, we decomposed and tracked 76 MUs from pre- to post-AIH (10.9 ± 7.71 per participant; range of 4-23), whereas we decomposed and tracked 75 MUs from pre- to post-SHAM (10.7 ± 6.82 per participant; range of 3-22).

### Maximal strength is enhanced by AIH

The force output increased from PRE to POST in the AIH condition but remained unchanged in the SHAM condition. The linear mixed effects model revealed a significant interaction between condition and timepoint (χ^2^(1) = 9.02, p = 0.003). Across muscles, force output increased significantly from PRE (73.3 ± 12.3N) to POST (114.3 ± 12.3N) in the AIH condition (p = 0.002) but did not change in the SHAM condition (PRE = 93.1 ± 12.3N; POST = 87.8 ± 12.3N; p = 0.999). This confirms our previous report that showed a significant increase in force output across muscles after AIH, but with a different statistical analysis (Afsharipour *et al*., 2022).

### Motor unit discharge rates are enhanced by AIH

MU discharge rates increased from PRE to POST in the AIH condition (see example in figure 1) but remained unchanged in the SHAM condition (see example in figure 2). The linear mixed effects model revealed a significant interaction between condition, timepoint, and muscle (χ^2^(1) = 9.19, p = 0.002). Mean discharge rates of BB MUs increased significantly (p < 0.0001) from PRE (16.7 ± 1.09pps) to POST (22.7 ± 1.06pps) in the AIH condition but remained unchanged across time in the SHAM condition (PRE = 16.8 ± 1.23pps; POST = 18.1 ± 1.22pps) p = 0.522). Similarly, mean discharge rates of TB MUs increased significantly (p = 0.002) from PRE (13.2 ± 1.12pps) to POST (15.2 ± 1.12pps) in the AIH condition but remained unchanged across time in the SHAM condition (12.9 ± 1.13pps; POST = 13.3 ± 1.15pps; p = 0.999). We have depicted a summary of these results in figure 3.

**Figure 1.**
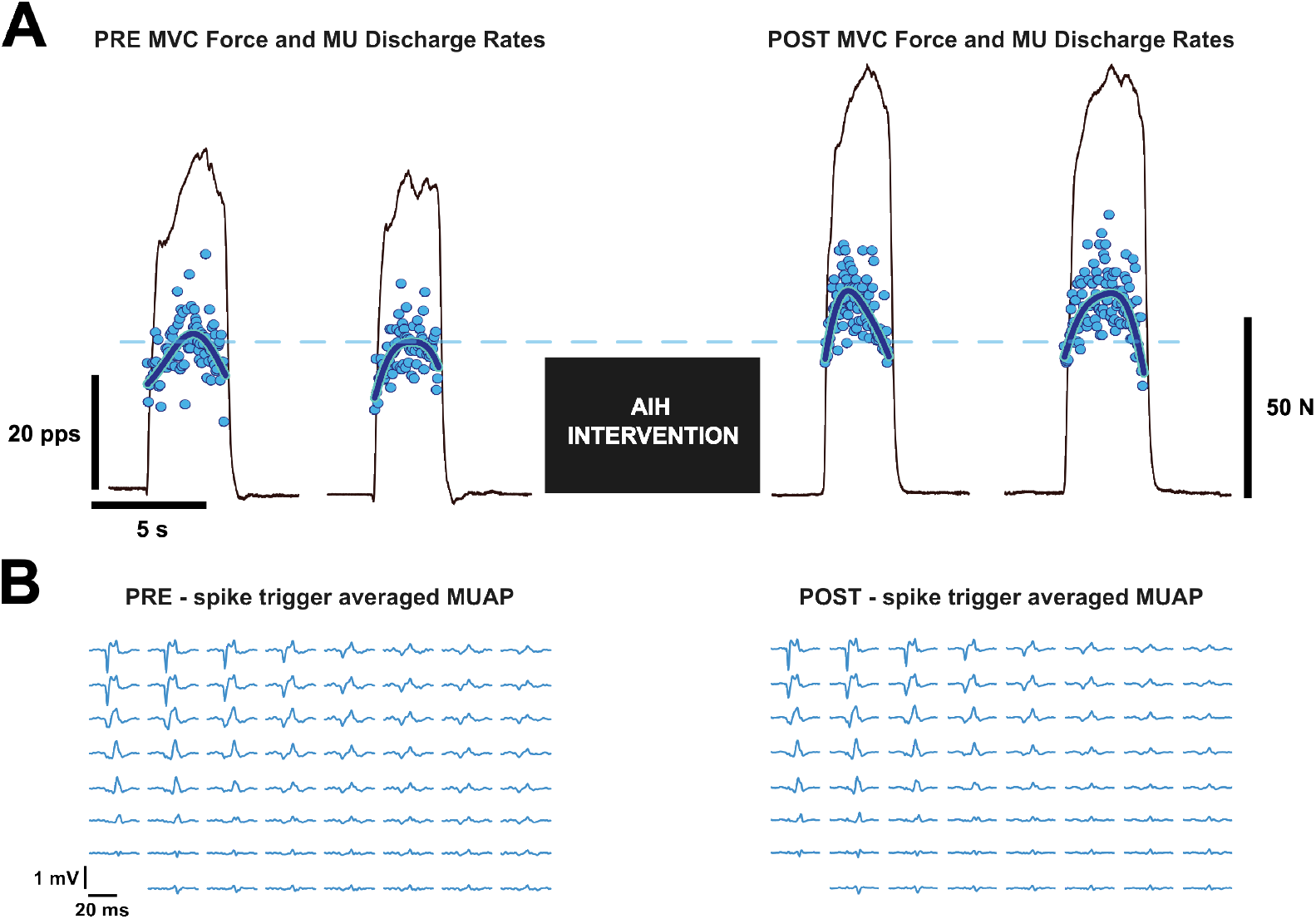
A: a single participant’s force (thin black line) from two maximal elbow flexion contractions before (left) and after (right) the acute intermittent hypoxia intervention. The instantaneous discharge rates (blue points) and smoothed discharge rate (thick black line) are also shown for an individual motor unit identified across both timepoints. The mean of the peak smoothed discharge rate from PRE is indicated by the dashed blue line for ease of comparison to the POST timepoint. B: the spike trigger averaged motor unit action potential identified across the electrode array. Note that the waveforms are similar across timepoints (left vs right), despite tracking using the filter transfer approach.

**Figure 2.**
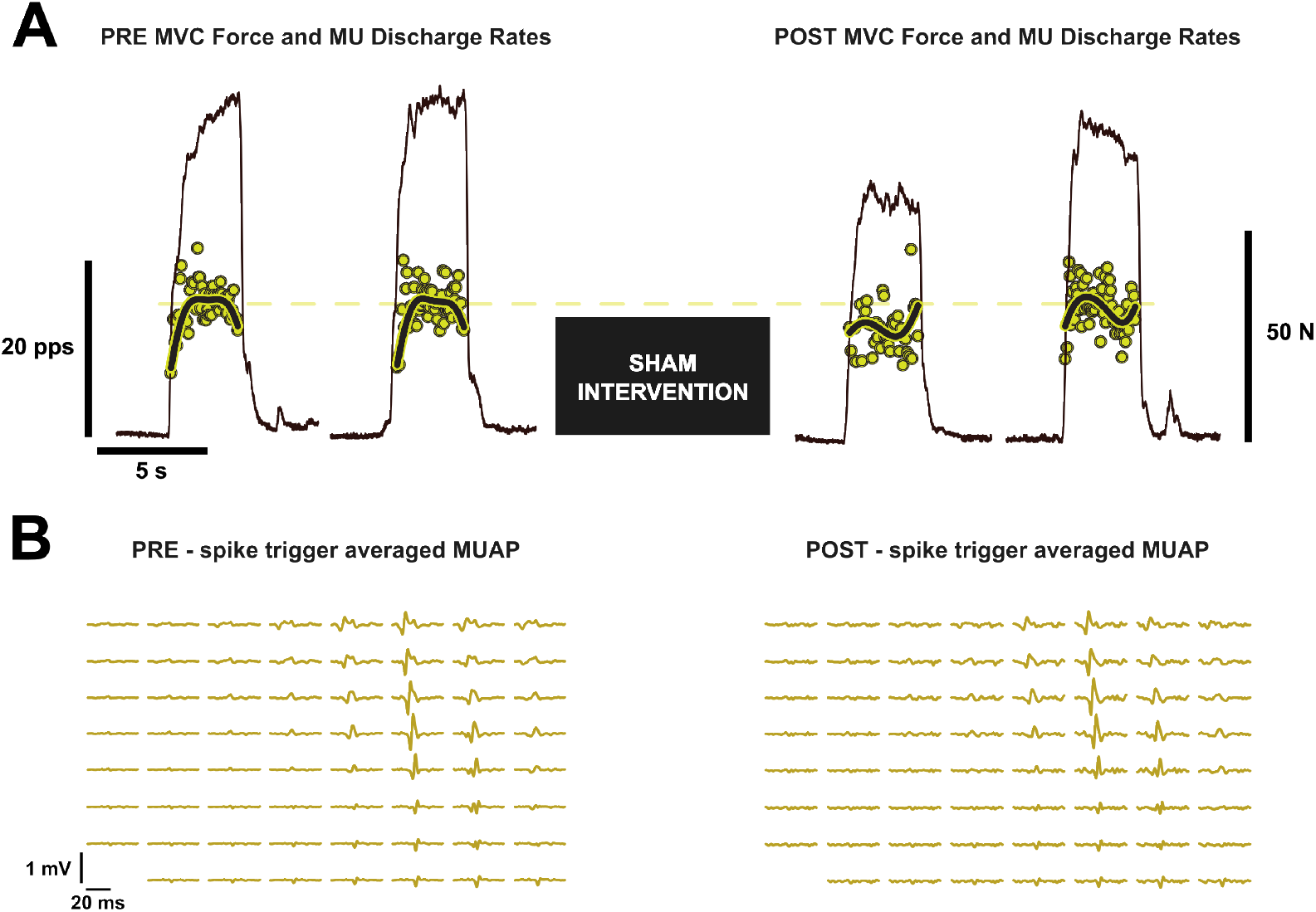
A: a single participant’s force (thin black line) from two maximal elbow flexion contractions before (left) and after (right) the sham intervention. The instantaneous discharge rates (yellow points) and smoothed discharge rate (thick black line) are also shown for an individual motor unit identified across both timepoints. The mean of the peak smoothed discharge rate from PRE is indicated by the dashed yellow line for ease of comparison to the POST timepoint. B: the spike trigger averaged motor unit action potential identified across the electrode array. Note that the waveforms are similar across timepoints (left vs right), despite tracking using the filter transfer approach.

**Figure 3.**
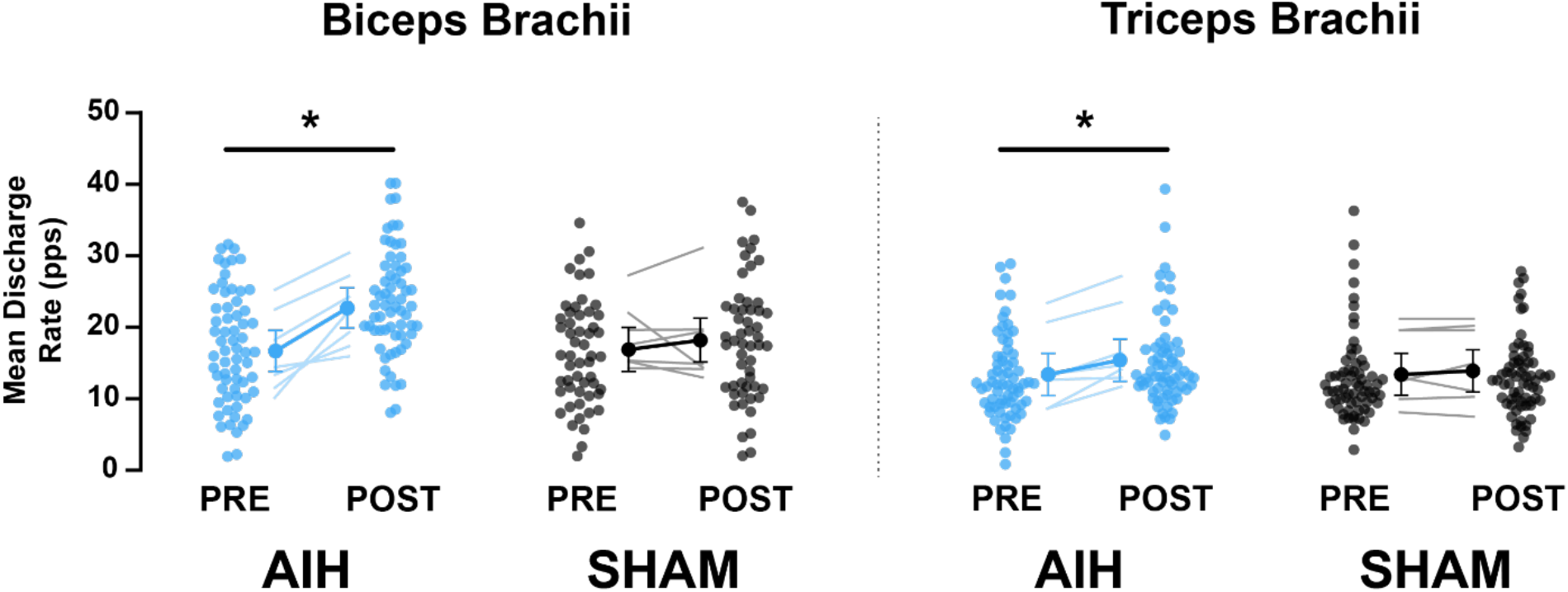
Mean discharge rates for biceps brachii (left panel) and triceps brachii (right panel) motor units during the AIH (blue) and SHAM (black) conditions. Individual motor units were tracked from PRE to POST in each condition and are indicated by a single data point. In the center of each panel, the mean change in discharge rate for each participant is indicated by a thin line, whereas the model effect is shown with the points and thick lines with error bars indicating the 95% confidence interval. Asterisks indicate a significant change from PRE to POST.

## DISCUSSION

The present study aimed to investigate the effects of a single session of AIH on MU discharge rates during *maximal* voluntary contractions in people with chronic incomplete spinal cord injury at the cervical level. Our novel results indicate that MU discharge rates in the BB and TB during *maximal* isometric elbow flexion and extension contractions, respectively, were substantially increased after AIH, but remained unaltered after SHAM AIH. We propose that AIH may present a novel approach for increasing spinal motoneuronal excitability and/or facilitating excitation to the motoneurons in the somatic nervous system of people with impaired force output of neural origin. The effects likely arise from a combination of direct and indirect serotonergic influences on motoneuron excitability and/or via enhanced synaptic transmission to the motoneurons.

Damage to descending motor tracts (i.e., corticospinal and brainstem motor pathways) resulting from spinal cord injuries can have devastating effects on excitatory and inhibitory ionotropic synaptic inputs (Oudega & Perez, 2012) and on descending neuromodulatory synaptic inputs (Holstege & Kuypers, 1987) to motoneurons below the level of injury. Combinations of excitation, inhibition and neuromodulation are the primary components of all motor commands in humans (Johnson *et al*., 2017; Khurram *et al*., 2022). Of particular importance after spinal cord injury, reduced excitatory input recruits fewer MUs, and causes them to discharge potentially at lower rates ((Thomas *et al*., 2014); *see below*). Therefore, increases in volitional muscle force after spinal cord injury can be achieved by either the recruitment of additional MUs, or by the enhanced discharge rates of previously recruited MUs. The current investigation provides support for the proposition that AIH causing an increase in maximal firing rates, but additional increases in motor unit recruitment remains a possibility.

In people with chronic incomplete spinal cord injury, both discharge rates during *maximal* force production, and the ability to recruit new MUs are impaired (Thomas *et al*., 2014). In terms of discharge rates, 18% (32/175) of identified triceps brachii MUs routinely discharged at <6 pps in people with chronic spinal cord injury during *maximal* voluntary contractions, whereas only 1/48 units in able-bodied participants discharged below 6 pps (Thomas *et al*., 1997). Discharge rates are also severely reduced in thenar MUs during *maximal* contractions in people with chronic incomplete spinal cord injury (rates ∼9 pps), compared to control subjects (rates ∼34 pps). These low discharge rates, however, are not due to an inability of a motoneuron to increase discharge but rather to reduced excitability or insufficient excitatory synaptic input to the motoneurons (Zijdewind *et al*., 2012). Zijdewind and colleagues (2012) showed that MU discharge rates could be increased when excitatory sensory input was applied during voluntary force outputs, such that small increases in discharge rates caused large alterations in force output, potentially due to the steepness of the force-frequency relationship (Thomas *et al*., 1991) at which the spinal cord injured MUs in the current study are discharging prior to AIH. Prior to AIH, as well as before and after the SHAM intervention, BB (∼17 pps) and TB (∼13 pps) discharge rates were lower than those we have typically observed in people without spinal cord injury (unpublished preliminary findings), however discharge rates increased by ∼6 pps and ∼2 pps in the BB and TB muscles, respectively, after the AIH intervention.

### Mechanisms responsible for increased discharge after AIH

The initial ideas for the use of AIH to enhance neuronal function were derived from studies of the phrenic motor output modulation that resulted from electrical stimulation of carotid chemoafferent neurons (Millhorn *et al*., 1980*a*), which was determined to be serotonin-dependent (Millhorn *et al*., 1980*b*). AIH, it turned out, had similar effects on phrenic motor output (Hayashi et al 1993), which led to several decades of work from Mitchell and colleagues (for reviews, see: (Dale-Nagle *et al*., 2010; Dale *et al*., 2014; Gonzalez-Rothi *et al*., 2015; Welch *et al*., 2020; Mitchell & Baker, 2022; Vose *et al*., 2022)) showing that AIH stimulates the carotid body chemoreceptors and causes the release of serotonin within the central nervous system(Baker-Herman & Mitchell, 2002; MacFarlane & Mitchell, 2009). After spinal cord injury, spinal motoneurons are highly sensitive to residual descending neurotransmitter release of serotonin (Bennett *et al*., 2001; Li & Bennett, 2003). This hyperexcitability is the result of constitutively active 5-HT_2c_ receptors (Li *et al*., 2007; Murray *et al*., 2010, 2011; Tysseling *et al*., 2017) and could render the AIH intervention, which causes the release of 5-HT from the brainstem via spared descending tracts, to have profound effects on motoneuron excitability.

Serotonergic neuromodulation is an important contributor for *maximal* voluntary activation of muscle in humans (for review see (Kavanagh & Taylor, 2022). Using pharmaceutical interventions in healthy young adults, Kavanagh and colleagues have shown that paroxetine (i.e., a selective serotonin reuptake inhibitor) increases *maximal* voluntary contraction strength while cyproheptadine (i.e., a competitive 5-HT_2_ receptor antagonist) decreases *maximal* voluntary contraction strength (Kavanagh *et al*., 2019; Thorstensen *et al*., 2021, 2022; Henderson *et al*., 2022). The same group also recently showed that ingestion of cyproheptadine reduces the rate of torque development and MU discharge rates during rapid contractions (Goodlich *et al*., 2022), and also reduces MU discharge rates and estimates of persistent inward currents during volitional isometric voluntary contractions (Goodlich *et al*., 2023). Taken together, these results suggest that serotonergic neuromodulation is important for voluntary force output in humans. If AIH does increase serotonin release onto motoneurons, facilitatory effects of 5-HT_2_ receptors on those motoneurons may explain the observed increases in MU discharge rates after AIH. Previous work in reduced preparations also shows a robust and complex pattern of activation within the rat spinal cord neurons both during and after an acute bout of hypoxia, providing further evidence for serotonergic modulation after AIH (Sandhu *et al*., 2015).

An alternative, or potentially complementary, explanation is that synaptic input to motoneurons is enhanced with AIH. Although damage to the spinal cord results in impaired descending motor commands, some connectivity of major descending motor tracts remains. Transcranial magnetic stimulation-evoked excitation and inhibition occurs in incomplete spinal cord injury, but the stimulator output required to evoke such responses in MU spike trains is greatly increased compared to people with intact spinal tracts (Smith *et al*., 2000). Christiansen and colleagues (2018) have shown that a single bout of AIH enhances corticospinal excitability, assessed with both transcranial magnetic stimulation and transcranial electrical stimulation, suggesting that the AIH-induced plasticity has a spinal origin. In a subsequent study, AIH also enhanced paired corticospinal-motoneuronal stimulation induced spinal synaptic plasticity in humans with chronic incomplete spinal cord injury (Christiansen *et al*., 2021). Such effects could be mediated by new BDNF protein synthesis, which then activates tropomyosin-related kinase B (Baker-Herman & Mitchell, 2002; Baker-Herman *et al*., 2004; Satriotomo *et al*., 2016; Dale *et al*., 2017). Further signaling cascades enhance the strength of excitatory synaptic inputs onto spinal motoneurons (Baker & Mitchell, 2000; Golder & Mitchell, 2005; Vinit *et al*., 2009; Lovett-Barr *et al*., 2012), all of which could enhance the excitatory synaptic input to motoneurons and increase the MU discharge rates as observed in the current investigation.

### Important Considerations

We must first consider the most important limitation in our study, which is the limited sample size of persons with iSCI, that was made difficult due to complications in recruitment that arose due to the COVID-19 global pandemic. Despite this low number of participants, we were highly successful in tracking several MUs from pre-to-post intervention (> 70 tracked MUs per muscle and condition).

Second, we did not assess the muscle activity of synergist muscles (i.e., brachialis or brachioradialis during flexion, or anconeus during extension). Thus, it is difficult to know whether AIH caused differences in load sharing among synergist muscles, which could have helped enhance net force output about the elbow.

Third, it is quite tempting to speculate about the effects of AIH on alterations in the recruitment thresholds of MUs, the disproportionate effects of AIH on low- vs higher-threshold MUs, and the recruitment of previously inactive MUs; but we must be conservative and acknowledge the limitations of our own experimental methods. To this end, we have avoided analysis involving recruitment thresholds because participants were not instructed to perform the ascending force in a controlled manner (i.e., the rate of force development was not constrained). This presents several issues: 1) faster contractions (i.e., rapid contractions) compress recruitment thresholds, and 2) during fast contractions, failing to decompose a single spike at the beginning of a contraction could inflate the recruitment threshold by as much as 60-100% of MVC.

Finally, with regards to the assessment of whether there are newly recruited MUs after AIH, it is difficult to determine whether these MUs were recruited after AIH due to physiological reasons, or whether they were identified due to changes in the complexity of the EMG. Likewise, if there were more MUs identified during PRE than POST, it would be tempting to conclude that there were more MUs recruited during PRE than POST. Since the transfer of MU filters from less complex to more complex HDsEMG signals is methodologically problematic (Frančič & Holobar, 2021), we could falsely “lose” MUs when transferring the filters from PRE to POST AIH. Further work is needed to ensure such considerations are fully elucidated in the future.

### Potential for clinical applications

Current therapeutic approaches for improving volitional muscle force in people with spinal cord injuries are quite limited, but combining AIH with other therapies is potentially a helpful strategy. Enhanced motoneuron function is sure to contribute to meaningful improvements in functional tasks, allowing people with spinal cord injuries to generate forces required to both perform tasks that would be otherwise difficult, and also to have the strength required to perform physical rehabilitation linked to such tasks. The increase in MU discharge rates after AIH may also have potential implications for muscle fatigue or endurance (Martinez-Valdes *et al*., 2020), which should be addressed in the future. Although we studied the acute effects of AIH on MU discharge rates, it remains possible that repeated AIH may have cumulative effects and further enhance motoneuron function due to the downstream BDNF effects and protein synthesis (Hassan *et al*., 2018).

## CONCLUSION

The present study examined the effects of a single session of AIH on MU discharge rates during *maximal* voluntary efforts in participants with chronic incomplete spinal cord injury at the cervical level. Similar to previous reports in phrenic motoneurons of rodent models, we show that AIH caused a significant increase in MU discharge rates in the elbow flexors and extensors during *maximal* efforts. When subjected to SHAM AIH, MU discharge rates remained unaltered from pre- to post-intervention. Increases in MU discharge rates likely arise from direct or indirect effects of serotonin on motoneuronal excitability, and/or improved synaptic efficacy of excitatory commands. Taken together, the results from the present study suggest that AIH is capable of enhancing the discharge rate of MUs in people with incomplete spinal cord injury, which provides mechanistic insights about the origins of improvements in motor function that have been observed as a result of this novel therapeutic intervention. This approach may be particularly beneficial for people with neurological impairments or injuries that result in reduced volitional forces due to compromised neural connectivity.

## Data Availability

All data produced in the present study are available upon reasonable request to the authors

## Acknowledgements

The authors would like to extend thanks to Andres Cardona for his assistance with data collection.

## Data availability statement

The data that support the findings of this study are available on request from the corresponding author.

## Author contributions

G.E.P.P., A.H., M.S.S., and W.Z.R. conceptualized and designed the research; B.A. and M.S.S. performed the experiments; G.E.P.P., and A.H. analysed the data; G.E.P.P. interpreted the results of experiments; G.E.P.P. prepared the figures; G.E.P.P. drafted the manuscript; G.E.P.P., B.A., A.H., M.S.S., and W.Z.R. revised and approved the final version of the manuscript.

